# Sensitivity of SARS-CoV-2 antigen-detecting rapid tests for Omicron variant

**DOI:** 10.1101/2021.12.18.21268018

**Authors:** Meriem Bekliz, Francisco Perez-Rodriguez, Olha Puhach, Kenneth Adea, Stéfane Marques Melancia, Stephanie Baggio, Anna-Rita Corvaglia, Frédérique Jacquerioz-Bausch, Catia Alvarez, Manel Essaidi-Laziosi, Camille Escadafal, Laurent Kaiser, Isabella Eckerle

## Abstract

**Background:** The emergence of each novel SARS-CoV-2 variants of concern (VOCs) requires investigation of its potential impact on the performance of diagnostic tests in use, including Antigen-detecting rapid diagnostic tests (Ag-RDT). Although anecdotal reports have been circulating that the newly emerged Omicron variant is in principle detectable by Ag-RDTs, few data on sensitivity are available.

**Methods:** We have performed 1) analytical sensitivity testing with cultured virus in eight Ag-RDTs and 2) retrospective testing in duplicates with clinical samples from vaccinated individuals with Omicron (n=18) or Delta (n=17) breakthrough infection on seven Ag-RDTs.

**Findings:** Overall, we have found large heterogenicity between Ag-RDTs for detecting Omicron. When using cultured virus, we observed a trend towards lower sensitivity for Omicron detection compared to earlier circulating SARS-CoV-2 and the other VOCs. When comparing performance for Delta and Omicron in a comparable set of clinical samples in seven Ag-RDTs, 124/252 (49.2%) of all test performed showed a positive result for Omicron compared to 156/238 (65.6%) for Delta samples. Sensitivity for both Omicron and Delta between Ag-RDTs was highly variable. Four out of seven Ag-RDTs showed significantly lower sensitivity (p<0.001) to detect Omicron when compared to Delta while three had comparable sensitivity to Delta.

**Interpretation:** Sensitivity for detecting Omicron is highly variable between Ag-RDTs, necessitating a careful consideration when using these tests to guide infection prevention measures. While analytical and retrospective testing may be a proxy and timely solution to generate performance data, it is not a replacement for clinical evaluations which are urgently needed. Biological and technical reasons for detection failure by some Ag-RDTs need to be further investigated.

**Funding:** This work was supported by the Swiss National Science Foundation (grant numbers 196383, 196644 and 198412), the Fondation Ancrage Bienfaisance du Groupe Pictet, the Fondation Privée des Hôpiteaux Universitaires de Genève and FIND, the global alliance for diagnostics.

## Introduction

The emergence of each novel SARS-CoV-2 variants of concern (VOCs) requires investigation of its potential impact on the performance of diagnostic tests in use. SARS-CoV-2 antigen-detecting rapid diagnostic tests (Ag-RDT) offer quick, cheap and laboratory-independent results at the point of care.^1^ Although sensitivity is lower compared to the gold standard method, RT-PCR, they enable reliable detection of high viral load samples associated with infectious virus presence, making them impactful public health tools.^2,3^ However, the majority of Ag-RDT validation studies were performed prior to the emergence of SARS-CoV-2 variants of concern (VOC).^4^

The VOC Omicron was first reported at the end of November from South Africa and is characterized by a high number of mutations compared to earlier circulating SARS-CoV-2.^5^ The majority of mutations are located in the protein of the gene coding for the Spike protein, and, according to preliminary data, are associated with considerable escape from neutralization by both disease- and vaccine derived antibodies, and probably also associated to lower vaccine effectiveness.^6,7, 8, 9, 10^ Current epidemiological data show that Omicron circulation is associated with a steep increase in case numbers as well as an increased risk of reinfection.^11^

Beyond the Spike mutations, Omicron also has also mutations in the nucleocapsid, which is the target protein of almost all Ag-RDTs. Two mutations found in Omicron are R203K and G204R that have been described already before Omicron in some SARS-CoV-2 sequences. They were linked to increased sub-genomic RNA and increased viral loads.^12-14^ In addition, a deletion (Del31-33) is found in the nucleocapsid of Omicron, as well as another mutation P13L. No information on a potential impact of these mutations on Ag-RDTs performance is available so far. Anecdotal reports showed positive detection of Omicron-confirmed patient samples by Ag-RDTs but few experimental data on Ag-RDT sensitivity for Omicron are available.

## Methods

### Virus isolates

All viruses were isolated from clinical samples. Isolates were grown in Vero-E6 cells as described previously.^15^ The Omicron variant was initially isolated on Vero-TMPRSS cells, then further passaged with a stock passage (p2) prepared on VeroE6. Vero TMPRSS were kindly received from National Institute for Biological Standards and Controls (NIBSC, Cat. Nr. 100978). The following mutations and deletion in the nucleocapsid were present in the original patients’ sequence as well as in the virus isolate of the passage used in this study: R203K, G204R, P13L, Del31-33.

### Clinical specimens

Nasopharyngeal swabs for diagnostics of SARS-CoV-2 by RT-PCR collected from symptomatic individuals in the outpatient testing center of the Geneva University Hospital were included in this study. Infection with SARS-CoV-2 was diagnosed by RT-PCR assay (Cobas 6800, Roche). All samples originate from the diagnostic unit of the virology laboratory of the hospital and were received for primary diagnosis of SARS-CoV-2. Remaining samples were stored at -80°C, usually on the same day or within 24h. All samples had one freeze-thaw cycle before inoculation on cell cultures for infectious virus and for viral RNA quantification, for the majority of specimens the Ag-RDT was performed at the same time. Due to logistical constraints, a subset of specimens had one additional freeze-thaw cycle for Ag-RDT testing only. All specimens were characterized by full genome sequencing for their infecting SARS-CoV-2 variant.

### Viral load quantification

Viral loads in each sample were determined by quantitative real-time reverse transcription PCR (RT-qPCR) using SuperScript™ III Platinum™ One-Step qRT-PCR Kit (Invitrogen) after thawing. RT-PCR for SARS-CoV-2 E gene and quantification of genome copy number was performed as described previously.^16^ Presence of infectious virus was determined by nucleocapsid staining for infectious foci in Vero TMPRSS 24h after inoculation with the patient sample as described previously ^17^.

### Ag-RDT performance

The 8 commercially available Ag-RDT products used in the study are summarized in **Table S1**.

#### Analytical testing with cultured virus

Each isolate has undergone serial dilutions at 1:2 in DMEM. For each variant, we started the dilutions with the same virus concentration at 1.72E+04 PFU/mL. All Ag-RDT assays were performed according to the manufacturers’ instructions except that viral dilutions were added to the buffer instead of a swab specimen. All dilutions used for validation additionally were tested and quantified by RT-PCR assay for SARS-CoV-2 RNA copy numbers/mL. For each serial dilution of each variant, 5 µl of dilution has been applied to the proprietary buffer and then applied to the Ag-RDT using only materials provided in the kit.

#### Performance testing with clinical specimens

For testing with clinical specimens, 5 µl of VTM of each specimen has been directly added to the proprietary buffer, and then applied to the Ag-RDT in duplicates under BSL3 conditions.^18^ Ag-RDT buffer without virus was used as a negative control. All Ag-RDT assays were read visually in duplicate. All visible bands were considered as a positive result. The entire study was performed under BSL-3 conditions.

### Statistics

We first compared whether Log_10_ SARS-CoV-2 copies, days post symptom onset, and presence of infectious disease were significantly different between the Delta (n=18) and Omicron (n=17) patients using simple linear and logistic regressions. We then tested whether the overall sensitivities and discordances differed between Delta and Omicron using proportion tests. Finally, we compared sensitivities for Delta (n=34) and Omicron (n=36) tests separately for each Ag-RDT. To take into account that each patient had two independent tests, we used mixed-effect logistic regressions with tests nested into patients. Data were analysed using R4.1.2.

### Ethical approval

Ethical approval for samples used in this study for virus isolation was waived by the local ethics committee of the Geneva University Hospitals (HUG) that approves the usage of anonymized leftover patient samples collected for diagnostic purposes in accordance with our institutional and national regulations. The part of the study using patient specimens linked to clinical data (retrospective testing) was approved by the Cantonal ethics committee (CCER Nr. 2021-01488). For this part, all study participants and/or their legal guardians provided informed consent.

## Results

### Analytical testing with cultured SARS-CoV-2 isolates

We have evaluated analytical sensitivity using cultured SARS-CoV-2 Omicron variant, in comparison to previous data obtained on isolates of the other VOCs (Alpha, Beta, Gamma and Delta) and an early-pandemic (pre-VOC) SARS-CoV-2 isolate (B.1.610) in eight Ag-RDTs. Data on early pandemic SARS-CoV-2, Alpha, Beta, Gamma and Delta have been published previously but were included here for comparison to Omicron^15,18^.

Eight Ag-RDTs were used: I) Panbio COVID-19 Ag Rapid test device (Abbott); II) Standard Q COVID-19 Ag (SD Biosensor/Roche); III) Sure Status (Premier Medical Corporation); IV) 2019-nCoV Antigen test (Wondfo); V) Beijng Tigsun Diagnostics Co. Ltd (Tigsun); VI) Onsite COVID-19 Ag Rapid Test (CTK Biotech); VII) ACON biotech (Flowflex) and VIII) NowCheck Covid-19 Ag test (Bionote). This list includes all three Ag-RDTs on the WHO Emergency Use Listing (WHO-EUL) and the other tests that are on the waiting list for WHO-EUL approval.

When assessing by infectious virus titers (PFU/mL) (**Fig 1A**), analytical sensitivity to detect Omicron was lower than for the other VOCs in most of the tests evaluated. Two tests showed a slightly higher sensitivity for Omicron than for Delta (Test V and VII), but for these tests, both Delta and Omicron showed lower detection sensitivity than the other VOCs and pre-VOC SARS-CoV-2. The same pattern of lowest sensitivity for Omicron compared to the other VOCS was confirmed when assessing RNA copy numbers (**Fig. 1B**). Significant heterogenicity was observed between different Ag-RDTs to detect Omicron.

**Figure 1.**
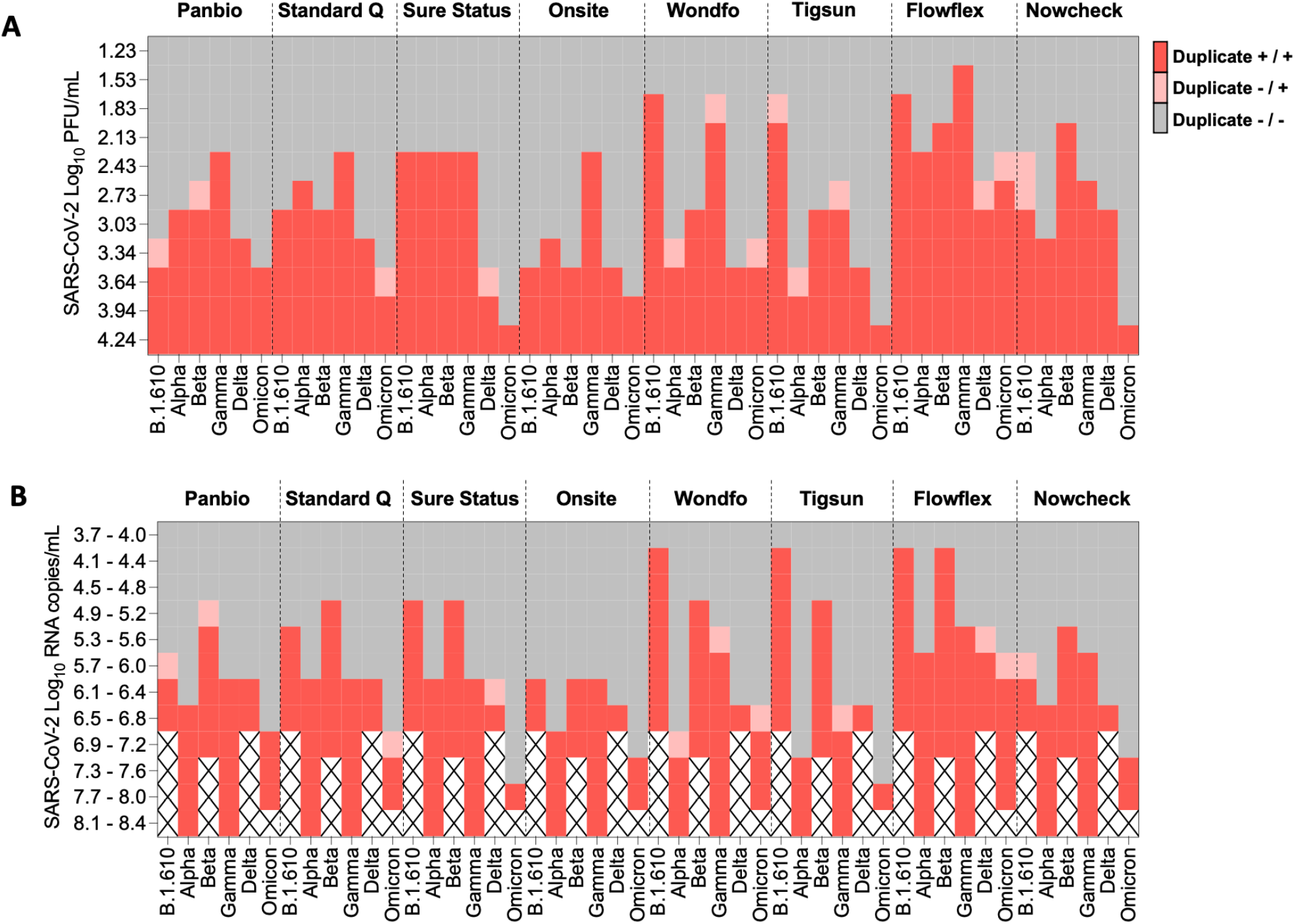
Heatmap based on Log_10_ PFU/mL (**Fig 1A**) and on RNA viral load ranges (**Fig 1B**) for analytical sensitivity of eight Ag-RDTs assays with an early-pandemic SARS-CoV-2 isolate (B.1.610), the VOCs Alpha, Beta, Gamma and Delta in comparison Omicron. Note: Analytical sensitivity for early-pandemic SARS-CoV-2 B.1.610, Alpha, Beta, Gamma and Delta have already been published before but were added here for consistency reasons and better interpretability of the data on Omicron.^15,16^

### Sensitivity testing in patient specimens

In addition to this analytical work, we have tested seven Ag-RDTs with original patient specimens as a retrospective sensitivity study with 35 nasopharyngeal specimens of confirmed Omicron (n=18) or Delta (n=17) breakthrough infections in vaccinated individuals during the first 5 days post-symptom onset. The two sample collections of Omicron and Delta patients’ specimens did not differ in RNA viral load, days post symptom onset or specimens with infectious virus presence (**Table 1**).

**Table 1.** Characteristics of clinical specimens. ^1^p-values for simple linear regressions (Log10 SARS-CoV-2 copies, DPOS) and simple logistic regression (Presence of infectious virus) are reported.

|  | Omicron (n=18) | Delta (n=17) | p<sup>1</sup> |
| --- | --- | --- | --- |
| Log10 SARS-CoV-2 copies, mean (SD) | 7.9 (0.7) | 8.0 (0.7) | .510 |
| DPOS, mean (SD) | 2.0 (1.2) | 1.9 (1.3) | .892 |
| Presence of infectious virus, n (%) | 14/18 (77.8%) | 14/17 (82.4%) | .613 |

**Table 2.** Detailed sensitivity for the seven Ag-RDTs tested with clinical samples. ^1^ p-values for logistic mixed-effect models (with tests nested into patients) are reported.

|  | Sensitivity (%) |  |  |
| --- | --- | --- | --- |
|  | Delta<br>(n=34) | Omicron<br>(n=36) | p<sup>1</sup> |
| Panbio | 67.7 | 36.1 | <.001 |
| Standard Q | 52.9 | 22.2 | <.001 |
| Sure Status | 52.9 | 27.8 | <.001 |
| Onsite | 64.7 | 47.2 | <.001 |
| Wondfo | 76.5 | 75.0 | .984 |
| Tigsun | 52.9 | 47.2 | .634 |
| Flowflex | 91.2 | 88.9 | .918 |

Testing with clinical specimens was done in duplicates for each specimens using seven Ag-RDTs to compare performance for Omicron and Delta infections (**Fig. 2**). When assessing overall test positivity, for Omicron 124/252 (49.2%) of tests showed a positive result compared to 156/238 (65.5%) (z = -3.65, p<.001). Of 126 test pairs, 14 showed a discordant result for Omicron vs. 7 in 119 test pairs performed for Delta (z = -1.46, p=.144). When comparing sensitivity for Delta vs. Omicron for each Ag-RDT, four Ag-RDTs showed significantly lower sensitivity (p<0.001) while three tests showed comparable performance (**Table 1 and Fig.3**). Sensitivity in our specimens panel ranged between 22.2% and 88.9% for Omicron and 52.9% to 91.2% for Delta, confirming the high variability of sensitivity between the different tests that was observed in our testing. The three tests that performed equally well had sensitivities between 47.2 and 91.2%.

**Figure 2.**
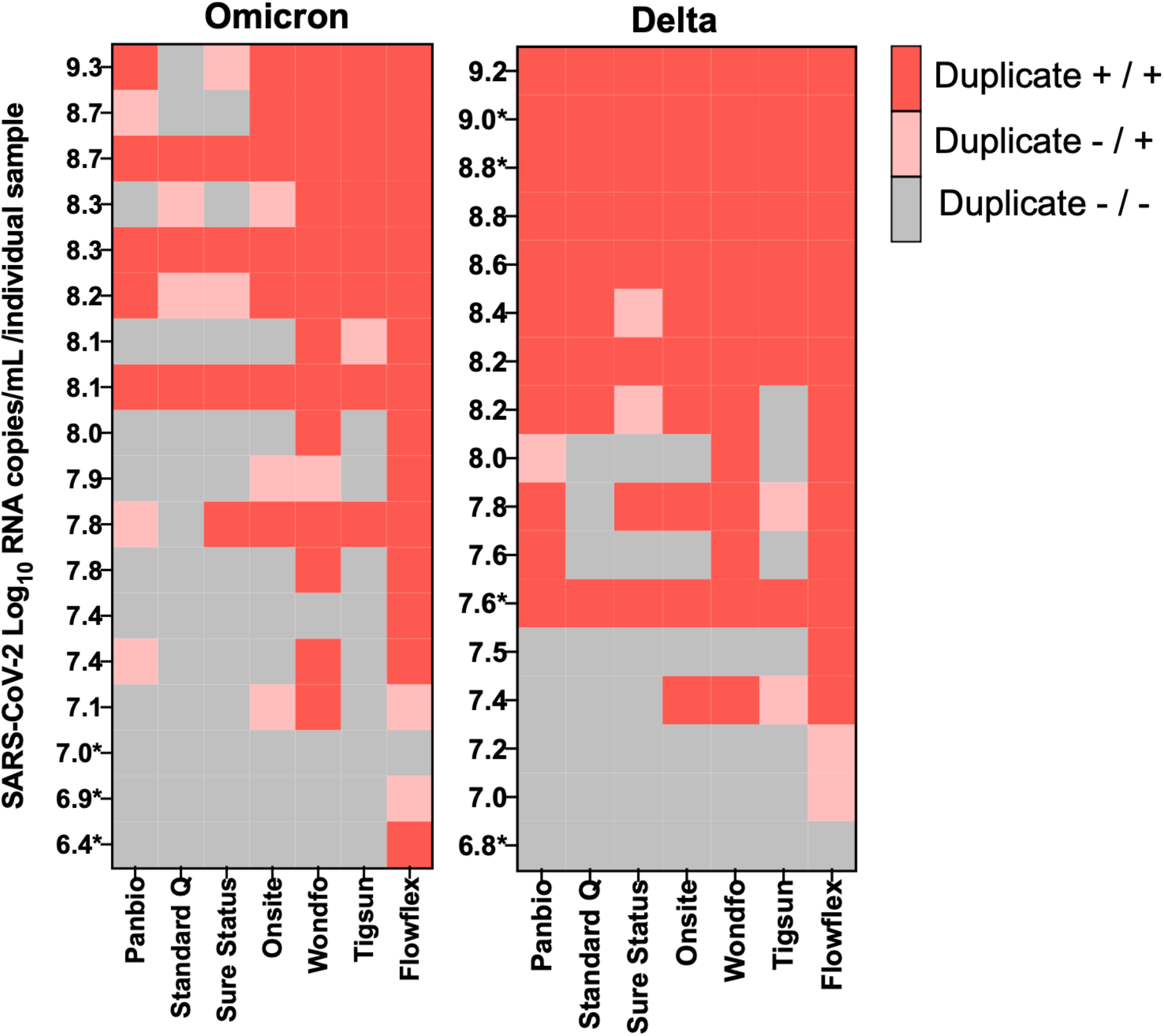
Heatmap of retrospective testing of original nasopharyngeal patient swab specimens from Omicron (n=18) and Delta (n=17) breakthrough infections in seven Ag-RDT assays per SARS-CoV-2 log_10_ RNA copies/mL, performed in duplicates. Infectious virus was detected from all patient specimens unless marked with * (* = no infectious virus isolated).

**Figure 3.**
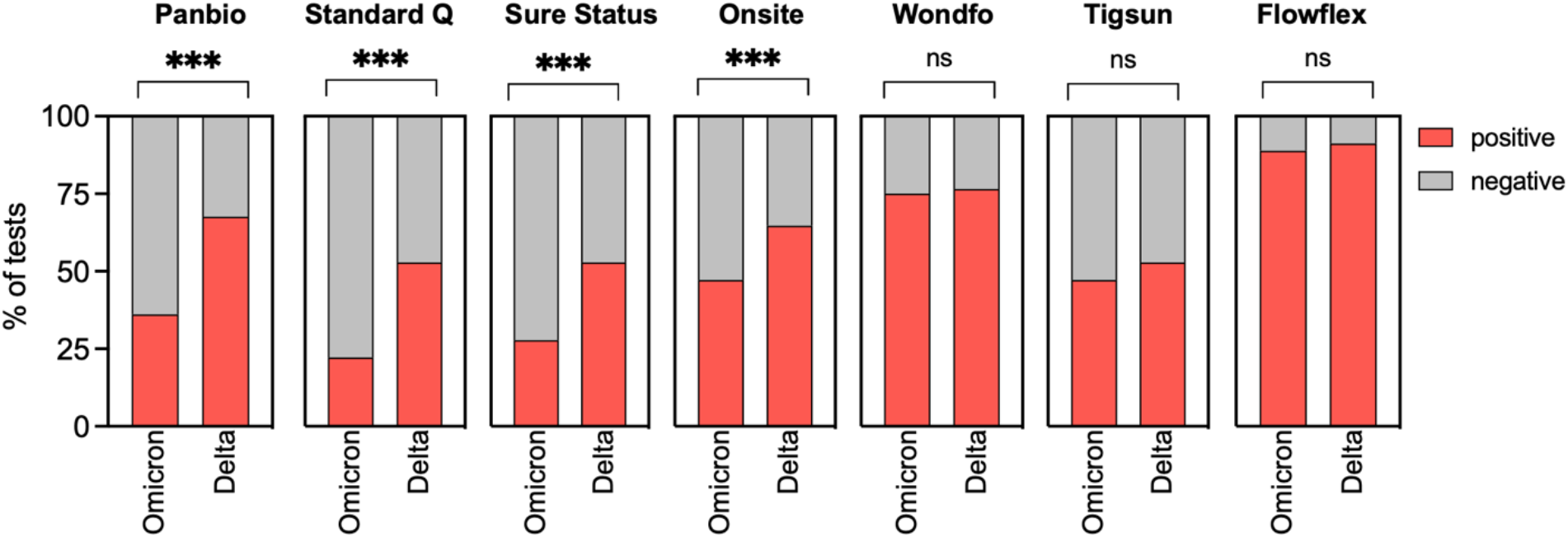
Percentage of positive/negative results for Omicron and Delta vaccine breakthrough infections per number of tests performed (Omicron n=36, Delta n=34). *** p<0.001, n.s., non-significant.

## Discussion

Newly emerging variants necessitate a rapid assessment of the performance of diagnostic tests in use. Here we have performed a comprehensive laboratory-based evaluation study of eight Ag-RDTs with cultured Omicron virus as well as a retrospective clinical validation with 35 patient specimens.

Overall, we have observed a lower sensitivity to cultured virus across different Ag-RDTs compared to earlier variants, suggesting that Omicron virus itself is detected with lower sensitivity than other variants. We have observed differences between Ag-RDTs from different manufacturers, but also between assessment for PFU and RNA copy numbers. Reasons are most likely due to different ratios between infectious particles and RNA copies among the different SARS-CoV-2 variants. Since the main public health benefit of Ag-RDTs are the detection individuals with infectious virus shedding and not just presence of viral RNA, assessment of infectious viral particles is of higher relevance in this context, and an overall tendency towards lower sensitivity was seen for both assessments. Of note, while in the analysis for infectious virus, the previous VOCs Alpha, Beta, Gamma and Delta were mainly detected with comparable or even higher sensitivity compared to pre-VOC SARS-CoV-2, and Omicron is the first VOC demonstrating a trend towards lower analytical sensitivity across assays.

Omicron has additional mutations in the nucleocapsid that have been previously observed in circulating SARS-CoV-2 before, although not largely present, in circulating SARS-CoV-2 before but so far their impact on Ag-RDT performance is unknown. The virus isolate used in our study carries all four of the known nucleocapsid mutations (P13L, Del31-33, R203K, G204R), confirmed from both patient specimens and virus isolate. Percentage of Omicron sequences with these mutations are 96.8% for P13L, 94.9% for Del31-33xx, 98.4 for R203K, and 98.4% for G204R of currently available Omicron sequences^19^. As not all circulating Omicron lineages harbour all mutations, additional analysis with such isolates would be of interest, however, at the time of conducting the study, no such isolates were available. However, our isolate represents the major circulating Omicron lineages.

In our clinical validation, we saw large heterogenicity between Ag-RDTs, with a loss of sensitivity for four Ag-RDT specimens. Comparisons of diagnostic assay by using different patient specimen collections are not trivial, and we have aimed for similar characteristics for the main determinants for rapid test performance, which is viral load, presence of infectious virus and time since days post symptom onset.^20,21^ Furthermore, we had access to detailed clinical data, and all specimens were from previously mRNA vaccinated individuals, followed by a Delta or Omicron breakthrough infection. At least in most high-income countries with high vaccination rates, this group of individuals is comprising the majority of Omicron infections observed, therefore our results are of immediate public health interest.

Few data are available so far on Ag-RDT performance for Omicron case detection. A small number of heterogeneous studies are available, but with little assessment for sensitivity and with conflicting results. A recent report from the U.S. Food & Drug Agency (FDA) announced that early data suggest reduced sensitivity for Omicron, in line with our findings, although no primary data are given.^22^ A study performed by Public Health England (PHE) with cultured isolated of Omicron and wild-type SARS-CoV-2 across dilutions ranging from 12.5 to 1250 focus forming units/mL and 30.000 to 4.070.000 viral copy numbers did not find a loss in sensitivity for five Ag-RDTs ^23^. Only one of the Ag-RDTs validated here, the Flowflex Ag-RDT, was also validated in our study. In our analytical testing, reduced sensitivity was seen for Omicron compared to wild-type SARS-CoV-2 in this test, but we did not see a difference in the clinical testing when compared to Delta. Overall, in both our assessments, this was the most sensitive Ag-RDT for most variants including Omicron. Another study used two nasal swab samples each from Omicron and Delta-infected individuals and validated the Abbott Binax Now Ag-RDT, a test that was not included in our study^24^ They conclude that Omicron can be detected by this test, although no extensive validation for sensitivity was performed. For the same test, data from a single clinical validation study are available from an outpatient testing Centre in the US using nasal swabs. ^25^ Sensitivity of a single antigen test was 95.2% for individuals with a cycle threshold value of the RT-PCR < 30, indicating good sensitivity with high viral load. A high failure rate was observed when oral specimens (cheek swabs) were used.

Strength of our study is that we have validated eight and seven Ag-RDT side-by-side for analytical and retrospective clinical sensitivity, respectively. Our selection of Ag-RDTs cover all of the three Ag-RDTs on the WHO-EUL, and three others that are on the WHO-EUL waiting list for approval, thus of high global public health relevance.^26,27^ If the lower sensitivity towards Omicron that we observed here is confirmed by findings from clinical validations at the point of care, the use of Ag-RDTs in the early symptomatic period of an Omicron infection or in asymptomatic patients could be less reliable, with possibly important implications for public health measures. However, all Ag-RDTs were able to detect Omicron infections and so far, there is no reason to change advice on how to implement RDTs to support testing and COVID response strategies. As our evaluation here was rather focused at the lower end of detection, results might be of higher relevance to testing in an asymptomatic population or in the very early infection phase, but not necessarily to the acute symptomatic infection phase when peak viral loads are reached.

Our study has several limitations. For cultured virus, the ratio between infectious virus, viral protein and RNA copies might differ considerably to original human specimens. The retrospective testing is done with only a low number of patients swab samples that have been submerged in viral transport medium, whereas the recommended sample type for Ag-RDT use is fresh swabs. This has introduced an extra dilution factor as well as an additional freeze/thaw cycle. Although we tried to reduce the number of freeze-thaw cycles to a minimum, we cannot exclude loss of RNA, protein or infectious virus, thus not reflecting fully the characteristics of a fresh patient specimen. To correct for loss of RNA after the first freeze-thaw cycle, we have re-tested viral RNA loads by RT-PCR and have used these values for comparison. Another limitation is that to compare across assays we have used the same approach as we did for analytical testing, with only 5 μL of the original patient VTM added to the buffer of each kit to be able to use the same specimens for testing with a high number of tests in parallel. The volume of viral transport medium added to the buffer was lower than what was recommended by some manufacturers, and for some Ag-RDTs there was no recommendation on the use of swab samples in VTM. Therefore, viral loads of the original sample and sensitivities observed in our sample collection cannot be compared to results obtained from clinical validations performed on fresh samples and our results should be interpreted as a comparison between Ag-RDTs and not as sensitivity thresholds for absolute viral loads and/or presence of infectious virus. Rather, we have investigated the lower end of sensitivity in the Ag-RDTs tested. Therefore, a reduced sensitivity in some tests, but not complete failure to detect Omicron could be of higher relevance in the beginning of the infection, when viral loads are still on the rise, and of less relevance once peak viral loads are reached.

Lower sensitivity observed in this study could be due to a variant-specific impact on Ag-RDT performance. However, since many Omicron infections are currently observed in vaccinated individuals, it remains unclear if virus shedding and test performance differs between unvaccinated and vaccinated individuals, and no studies are available investigating Ag-RDT performance in unvaccinated vs. vaccinated individuals are available yet. To date, most validation studies of Ag-RDTs were done in the first year of the pandemic, before circulation of VOCs and in mostly immune-naïve individuals experiencing their primary SARS-CoV-2 infection. Other factors, such as *in vivo* shedding of infectious virus and overall viral can be one reason for differences in test performance. However, we have shown recently that neither RNA viral loads nor infectious titers differ significantly between Omicron and Delta breakthrough infections, thus differences in viral load are unlikely the reason for lower sensitivity in Omicron in some tests.^17^

Importantly, while analytical and retrospective testing may be a proxy for clinical sensitivity, is not a replacement for clinical evaluations at the point of care. The discrepancies in our results between testing with cultured virus and retrospective patient samples highlights the need for proper clinical studies in well-defined patient cohorts. Therefore, further studies on diagnostic accuracy of Ag-RDTs performed at the point of care for the newly emerged VOC Omicron are urgently needed to guide public health responses.

## Data Availability

All data produced in the present study are available upon reasonable request to the authors

## Acknowledgments

We thank the patients for participating in our study. We thank Pauline Vetter for help with clinical data. We thank Silvio Steiner, Jenna Kelly and Volker Thiel for sequencing of the Omicron isolate.

## Supplementary material

### Tables

**Table S1.** Overview of Ag-RDTs kits evaluated in the study.

|  | **Name of kit** | **Manufacturer** | **Target protein** |
| --- | --- | --- | --- |
| I | Panbio, COVID-19 Ag Rapid test device | Abbott | Nucleocapsid |
| II | Standard Q COVID-19 Ag | SD BIOSENSOR (Roche) | Nucleocapsid |
| III | Sure Status | Premier Medical Corporation | Nucleocapsid |
| IV | 2019-nCoV Antigen test | Wondfo | Nucleocapsid |
| V | Beijng Tigsun Diagnostics Co. Ltd | Tigsun | Nucleocapsid |
| VI | CTK biotech | Onsite | Nucleocapsid |
| VII | ACON biotech | Flowflex | Nucleocapsid |
| VIII | NowCheck Covid- 19 Ag test | Bionote | Nucleocapsid |

